# Therapeutic use of cannabis, cannabis-derived products and synthetic cannabinoids for rheumatoid arthritis: Protocol for a systematic review

**DOI:** 10.1101/2020.07.25.20149922

**Authors:** Clara Schulze, Josefina Durán, Rocío Bravo-Jeria, Francisca Verdugo-Paiva, Macarena Morel, Gabriel Rada

**Affiliations:** Faculty of Medicine, Pontificia Universidad Católica de Chile, Santiago, Chile; Epistemonikos Foundation, Santiago, Chile; UC Evidence Center, Cochrane Chile Associated Center, Pontificia Universidad Católica de Chile, Santiago, Chile; Internal Medicine Department, Faculty of Medicine, Pontificia Universidad Católica de Chile, Santiago, Chile; Clinical Immunology and Rheumatology Department, Faculty of Medicine, Pontificia Universidad Católica de Chile, Santiago, Chile

**Keywords:** cannabis, cannabis-derived products, marijuana, cannabinoids, synthetic cannabinoids, medical cannabis, rheumatoid arthritis, systematic review, meta-analysis

## Abstract

**OBJECTIVE:** To assess the therapeutic effects of cannabis, cannabis-derived products and synthetic cannabinoids for rheumatoid arthritis.

**DESIGN:** This is the protocol of a systematic review. DATA SOURCES: Searches will be conducted in PubMed/Medline, Embase, Cochrane Central Register of Controlled Trials (CENTRAL), trial registries, grey literature and in a centralized repository in L-OVE (Living OVerview of Evidence). L-OVE is a platform that maps PICO questions to evidence from Epistemonikos database.

**ELIGIBILITY CRITERIA FOR SELECTING STUDIES AND METHODS:** We will include randomized controlled trials evaluating therapeutic use of cannabis, cannabis-derived products and synthetic cannabinoids for rheumatoid arthritis. Our primary interest will be in trials comparing the intervention with placebo or no treatment (intervention plus optimal treatment vs placebo plus optimal treatment or optimal treatment alone) in patients receiving optimal treatment for rheumatoid arthritis. Optimal treatment will be defined as disease modifying anti-rheumatic drugs. Two reviewers will independently screen each study for eligibility, extract data, and assess the risk of bias. We will perform random-effects meta-analyses and use Grading of Recommendations Assessment, Development and Evaluation (GRADE) to assess the certainty of the evidence for each outcome.

**ETHICS AND DISSEMINATION:** The Scientific Ethics Committee of the Pontificia Universidad Católica de Chile granted ethical exemption for the realization of this study. Results of this review will be widely disseminated via peer-reviewed publications, social networks and traditional media and will be sent to relevant international organizations discussing this topic.

## INTRODUCTION

Rheumatoid arthritis is a chronic systemic auto-immune disorder of unknown etiology characterized by persistent synovitis, systemic inflammation, and autoantibodies (1), which leads to joint damage (2,3) and extra-articular manifestations (such as cardio and cerebrovascular disease, depression and cognitive impairment (4–11). It is the most common inflammatory arthropathy (6), with a higher prevalence among women and adults around the sixth decade of life (12,13).

There is no cure for rheumatoid arthritis (3). Disease modifying anti-rheumatic drugs (DMARDs), non-steroidal anti-inflammatory drugs and steroids are medicines from rheumatoid arthritis therapeutic arsenal (3). Although clinical remission or low disease activity are achievable goals, symptoms and disease burden still leads to disability, reduced work productivity, poorer quality of life related to pain and disease activity (12,14–18) and significant excess mortality (19).

Cannabis is a generic term used for drugs derived from plants belonging to the genus cannabis, mainly from two species: Cannabis sativa and Cannabis indica (20), with Δ-9-tetrahydrocannabinol and cannabidiol (CBD) as major constituents (21). Cannabinoids is the term to refer to the ligands of cannabinoid receptors (22). Even though there is an overlap between categories, cannabinoids are usually clustered in three groups: endocannabinoids (produced by the body), plant cannabinoids or phytocannabinoids (produced by cannabis plants, or other plants [e.g. Echinacea]) and synthetic cannabinoids (23).

Medical use of cannabis, cannabis-derived products and synthetic cannabinoids for rheumatoid arthritis is partially substantiated by laboratory and animal models suggesting they could reduce inflammatory cell infiltration, slow bone erosion and reduce disease severity (24–27). However, two systematic reviews addressing cannabinoids revealed that common short-term adverse events in humans include immediate psychomotor effects, dizziness, nausea, xerostomia and tachycardia (28,29); rare serious adverse effects include stroke, seizures, myocardial infarction, acute kidney injury, and psychosis (28,29) and long term risks associated with medical cannabis use in patients with rheumatoid arthritis remain unknown.

Even though the mechanism of action of cannabis, cannabis-derived products and synthetic cannabinoids in rheumatoid arthritis is far from being completely elucidated, and results to date have been disappointing, the widespread use and popular belief about the therapeutic effects of cannabis has fostered the conduction of many systematic reviews that encompass or address in some way our clinical question (30–37). However, since their publication, not only the body of evidence has grown but also the list of available synthetic cannabinoids.

This systematic review aims to contribute to the benefit/risk ratio judgment of cannabis, cannabis-derived products and synthetic cannabinoids medical use in rheumatoid arthritis, due to public therapeutic use support in the absence of a strong evidence base, rheumatoid arthritis patients’ increasing inquiring in their use and clinicians concerns about their risks (38,39). Therefore, recent pieces of evidence not addressed by previous reviews must be included and more exhaustive and sensitive searches seem to be necessary to encompass terminological variety associated with cannabis, cannabis-derived products and synthetic cannabinoids.

## OBJECTIVE

To assess the therapeutic effects of cannabis, cannabis-derived products and synthetic cannabinoids for rheumatoid arthritis.

## METHODS

### PROTOCOL AND REGISTRATION

This review is part of a larger project of multiple parallel systematic reviews for any condition for which cannabis, cannabis-derived products or synthetic cannabinoids have been postulated as a therapeutic alternative. The protocol of the larger project is registered on PROSPERO and was allocated the registration number CRD42018097382 (40). The protocol was adapted to the specificities of the question assessed in this review in line with the Preferred Reporting Items for Systematic Reviews and Meta-Analyses Protocols (PRISMA-P) (41) and submitted to PROSPERO (awaiting ID allocation). The PRISMA-P checklist is given as supplement (see Supplement 1: PRISMA-P checklist archive).

### AMENDMENTS

In the event that an amendment to this protocol is necessary, we will record the date of each amendment along with a description of the change and the rationale.

### ELIGIBILITY CRITERIA

#### Study designs

We will only include randomized controlled trials (RCTs). There will be no restrictions by publication status (i.e. published, unpublished, in press, in progress).

#### Participants

We will include trials involving participants of any age with rheumatoid arthritis, with clinical diagnosis or who met the rheumatoid arthritis criteria of the 1987 American College of Rheumatology (ACR) Classification criteria for rheumatoid arthritis (42) or the 2009-2010 ACR/European League Against Rheumatism (EULAR) Classification criteria for rheumatoid arthritis (43), whichever was used by study authors. If we find substantial clinical heterogeneity on how the condition was defined we will explore this using sensitivity analysis.

Studies involving participants with conditions other than rheumatoid arthritis (i.e. mixed populations) will be included only if outcomes for participants with rheumatoid arthritis are presented as a separate data subset or if separate data is available from the study authors.

Studies evaluating the effects on people with idiopathic juvenile arthritis, other inflammatory arthritis, healthy volunteers, animal models or in vitro conditions will be excluded.

#### Interventions

In order to have a comprehensive picture of the role of interventions exerting action in the endocannabinoid system, the interventions of interest will be cannabis, any product derived from cannabis and synthetic cannabinoids.

Trials evaluating other non-processed plants containing cannabinoids (e.g. Echinacea) will be excluded, but preparations that extract cannabinoids from these plants will be included.

Any drug stimulating CB1 or CB2 receptors (e.g. agonists, drugs that increase the level of endocannabinoids by blocking degradation) will be included. Antagonists of cannabinoids receptors will be excluded. We will not restrict by route of administration or dose.

Some examples of interventions that will be included are:

- Herbal cannabis: Marijuana, hashish.
- Cannabis-derived products: Nabiximol (e.g. Sativex®), dronabinol (e.g. Marinol®), CBD.
- Synthetic cannabinoids: Cannabinoid receptor agonists (e.g. Levonantradol, Nabilone, lenabasum), fatty acid amide hydrolase inhibitors and inactivators.

#### Comparators

Our primary interest will be in trials evaluating the additive effect of cannabis, cannabis-derived products or synthetic cannabinoids in patients receiving optimal treatment for rheumatoid arthritis versus placebo or no treatment (intervention plus optimal treatment vs placebo plus optimal treatment or optimal treatment alone). Optimal treatment will be defined as DMARDs (e.g. methotrexate, leflunomide, abatacept, adalimumab).

However, we will include any comparison, organized in the following categories:

Primary comparisons:

- Intervention plus optimal treatment versus placebo plus optimal treatment.
- Intervention plus optimal treatment versus optimal treatment.

Secondary comparisons:

- Intervention versus placebo.
- Intervention plus non-standard treatment versus non-standard treatment.
- Comparison between different interventions, routes of administration or doses.

#### Outcomes

We used the core outcome set developed by the World Health Organization (WHO) and International League of Associations for Rheumatology (ILAR) (WHO-ILAR) (44), previously published guidelines and reviews, and the judgment of the authors of this review in order to select the primary and secondary outcomes, as well as to decide upon inclusion.

Primary outcomes:

- Disease activity.
- Pain.
- Physical disability.

Secondary outcomes:

- Quality of life.
- Serious adverse events (SAEs).
- Nervous system adverse events.
- Psychiatric adverse events.

Other outcomes:

- Tender joints.
- Swollen joints.
- Physician global assessment.
- Patient global assessment.
- Acute phase reactants.
- Mortality.
- Sleep.
- Depression.
- Fatigue.
- Anxiety.
- Psychological distress.

We will prioritize primary and secondary outcomes for the development of the GRADE ‘Summary of Findings’ table (45). A table with all other outcomes will be presented as an appendix.

Outcomes will not be considered part of the eligibility criteria. Any article meeting all the criteria except for the outcome criterion will be preliminarily included and evaluated in full text.

#### Timing

We will consider grouping outcomes according to the time point in which they were measured in categories (e.g. short term, medium term, long term). We will prioritize long term outcomes, especially in outcomes related to disease activity.

#### Setting

There will be no restrictions by type of setting.

#### Language

There will be no language restrictions.

## SEARCH METHODS FOR IDENTIFICATION OF STUDIES

### ELECTRONIC SEARCHES

We will conduct sensitive electronic searches of the following databases (without any date, publication status or language restriction):

- Cochrane Central Register of Controlled Trials (Wiley).
- MEDLINE (PubMed).
- EMBASE (Elsevier).

The MEDLINE strategy was adapted to the syntax and subject headings of the other databases (see Supplement 2: Search strategy for electronic databases). The search will cover the period until the day before submission to a journal and will be will be updated toward the end of the review.

### SEARCHING OTHER SOURCES

An expanded search will be performed to identify articles potentially missed through the database searches and in order to identify ‘grey literature’ and unpublished studies. This includes the following:

1. We will manually search reference lists of all included studies.
2. We will screen the reference lists of selected guidelines and narrative reviews on the topic.
3. We will conduct searches in Epistemonikos database (www.epistemonikos.org) and L·OVE (https://iloveevidence.com/) to identify other systematic reviews on the topic, scan their reference lists and evaluate in full text all the articles they include.
4. We will review Medwave (46), a peer-reviewed international general medical journal, in order to find FRISBEEs (FRIendly Summary of the Body of Evidence using Epistemonikos) that address our clinical question. FRISBEEs are rapid evidence summaries created from an evidence matrix in Epistemonikos (47). The evidence matrix is meta-analysed using the included primary studies and a summary of findings table is created using the GRADE method. Finally, the FRISBEE is drafted, peer-reviewed and published in Medwave (47).
5. We will create a matrix of evidence in Epistemonikos with all the existing systematic reviews identified, in order to identify other reviews that share primary studies with the index reviews.
6. We will conduct a cross-citation search in Google Scholar and Microsoft Academic, using each included study as the index reference.
7. We will hand search journals specialized in cannabis medicine: Cannabis, Cannabis and Cannabinoid Research.
8. We will review international conferences related to cannabis medicine: Scientific Meeting of the Research Society on Marijuana, International Cannabinoid Research Society Symposium on the Cannabinoids and Cann.
9. We will screen international conferences related to rheumatoid arthritis: ACR (https://acrabstracts.org/) and EULAR (https://www.eular.org/).
10. We will review websites from pharmaceutical companies producing cannabis-based products and other selected sites such as GW pharmaceuticals (https://www.gwpharm.com/), International Association for Cannabinoid Medicines (https://www.cannabis-med.org/?lng=en).
11. We will search the websites of regulatory agencies: USA Food and Drug Administration-MedWatch (https://www.fda.gov/safety/medwatch-fda-safety-information-and-adverse-event-reporting-program), European Medicines Evaluation Agency (https://www.ema.europa.eu/en), Australian Adverse Drug Reactions Bulletin (https://www.tga.gov.au/adr/aadrb.htm), and United Kingdom Medicines and Healthcare products Regulatory Agency pharmacovigilance and drug safety updates (https://www.gov.uk/government/organisations/medicines-and-healthcare-products-regulatory-agency).
12. The WHO International Clinical Trials Registry Platform Search Portal (http://apps.who.int/trialsearch/), the European Union Clinical Trials Register (https://www.clinicaltrialsregister.eu/), the United States National Institutes of Health Ongoing Trials Register (https://clinicaltrials.gov/), the ISRCTN registry (https://www.isrctn.com/), Current Controlled Trials (http://www.controlled-trials.com) and Natural Medicines (https://naturalmedicines.therapeuticresearch.com) will be searched to identify additional published or unpublished data and ongoing trials.
13. We will email the contact authors of all of the included trials to ask for additional publications or data on their studies, and for other studies in the topic.

## DATA COLLECTION AND ANALYSES

### STUDY SELECTION

The results of the literature search will be uploaded to the software CollaboratronTM (48). References will be de-duplicated by an algorithm comparing unique identifiers (database ID, DOI, trial registry ID), and citation details (i.e. author names, journal, year of publication, volume, number, pages, article title and article abstract). We will not anonymize the studies in any way before assessment.

At least two authors will independently screen each title and abstract identified by the search to decide upon their potential inclusion. We will obtain the full reports for all titles that appear to meet the inclusion criteria or require further analysis to decide on their inclusion. Then, full text articles will be screened against the inclusion criteria by at least two independent reviewers.

We will resolve disagreements through discussion or through a third reviewer if the discrepancy could not be solved. The reasons for excluding trials in any stage of the search will be recorded. We will outline the study selection process in a PRISMA flow diagram (see Figure 1).

**Figure 1.**
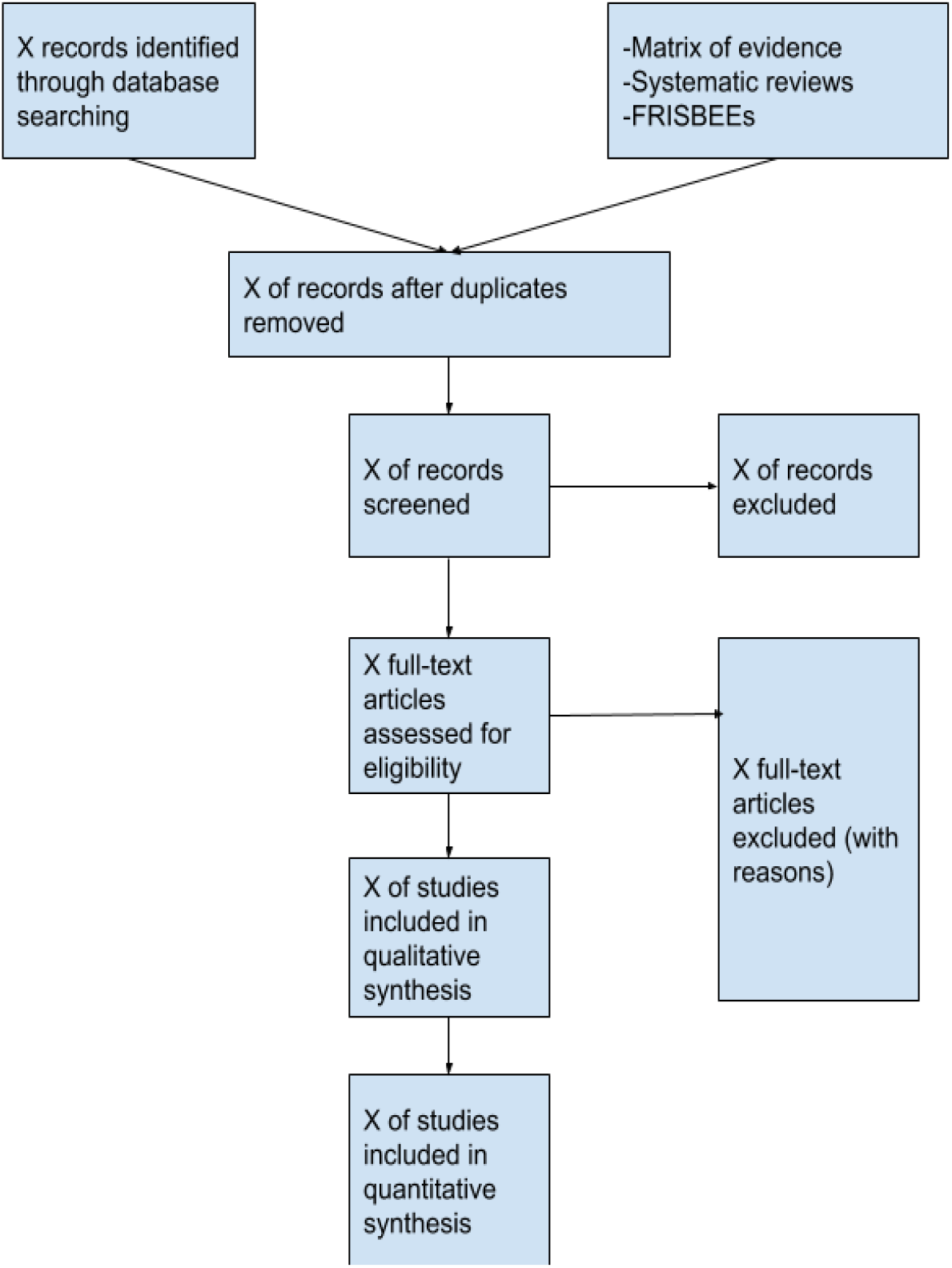
Preferred Reporting Items for Systematic Reviews and Meta-Analyses (PRISMA) flow diagram— A schematic of the processes of the systemic review. FRISBEEs: FRIendly Summary of the Body of Evidence using Epistemonikos.

### DATA EXTRACTION AND MANAGEMENT

Using standardized forms, two reviewers will extract data independently from each included trial. The following data will be extracted, where possible:

- Study design.
- Study eligibility criteria.
- Length of follow-up.
- Sample size.
- Rheumatoid arthritis diagnosis (clinical diagnosis or diagnosis using classification criteria such as ACR (42) or ACR/EULAR (43)).
- Characteristics of study population, including age, female/male ratio, geographic region, socioeconomic status (e.g., education and income), smoking habit, nutritional status, comorbidities (e.g. cardiovascular disease, malignancies, infections, gastrointestinal disease, osteoporosis, depression), disease duration (early versus established or years of evolution), severity and activity, structural damage (erosive rheumatoid arthritis) and previous treatments
- Details of the administered intervention(s) including dose, therapeutic scheme, route of administration and treatment duration.
- Details of any co-interventions, including current synthetic DMARDS, biologic DMARDS and steroids use.
- Method used to seek adverse reactions.
- Outcomes and timing of outcome assessment.
- Losses to follow up, exclusions and their reasons.
- Source of study funding.
- Conflicts of interest disclosed by the investigators.
- Risk of bias assessment for each individual study.

The raw data will be extracted for outcomes of interest. Whenever possible, we will use results from intention-to-treat analyses.

Differences in data extraction will be resolved by referring back to the original articles and establishing a consensus. One arbiter will adjudicate unresolved disagreements.

We will contact study authors to resolve any uncertainties.

### ASSESSMENT OF RISK OF BIAS IN INCLUDED STUDIES

At least two reviewers independently will assess risk of bias using a ‘risk of bias’ tool (RoB 2.0: a revised tool to assess risk of bias in randomized trials) (49). We will consider the effect of assignment to the intervention for this review. Two reviewers will independently evaluate five domains of bias for each outcome result of all reported outcomes and time points (bias due to the randomization process, deviations from intended interventions (effects of assignment to interventions at baseline), missing outcome data, and measurement of the outcome and selection of reported results). Answers to signaling questions and supporting information collectively will lead to a domain-level judgment in the form of ‘Low risk of bias’, ‘Some concerns’, or ‘High risk of bias’. These domain-level judgments will inform an overall ‘risk of bias’ judgment for each result. In case reported details in a study were insufficient, the original study investigators will be contacted for more information. Disagreements will be resolved first by discussion and then by consulting a third author for arbitration.

We will compute graphic representations of potential bias within and across studies using RevMan 5.3.5 (50).

### MEASURES OF TREATMENT EFFECTS

For dichotomous outcomes, we will express the estimate of treatment effect of an intervention as risk ratios (RR) or odds ratios (OR) together with 95% confidence intervals (CIs). If the outcome is a rare event (approximately less than 10%), we will use Peto odds ratio (Peto OR) instead of RR.

For continuous outcomes, we will use mean difference and standard deviation (SD) to summarize the data along with 95% CIs. Whenever continuous outcomes were measured using different scales, the treatment effect will be expressed as a standardized mean difference (SMD) with 95% CI. When possible, we will multiply the SMD by a standard deviation that is representative from the pooled studies, for example, the SD from a well-known scale used by several of the studies included in the analysis on which the result is based. In cases where the minimally important difference (MID) is known, we will also present continuous outcomes as MID units or inform the results as the difference in the proportion of patients achieving a minimal important effect between intervention and control (45). Then, these results will be displayed on the ‘Summary of Findings Table’ as mean difference (45).

### DEALING WITH MISSING DATA

When possible, we will contact the original authors of the study to obtain any missing data. If important numerical missing data could not be obtained, an imputation method will be used (51). When available, we will extract data from graphs by using data extraction software.

### ASSESSMENT OF HETEROGENEITY

We will assess the variations in treatment effect from the different trials by means of the I2 statistic. A rough guide to the interpretation of the I2 statistic given in the Cochrane Handbook is: 0–40% might not be important, 30–60% may represent moderate heterogeneity, 50–90% may represent substantial heterogeneity and 75–100% considerable heterogeneity. However, its importance will depend on magnitude and direction of effects and strength of evidence for heterogeneity (52).

### ASSESSMENT OF META-BIASES

We will investigate the presence of publication bias visually with the use of funnel plots. We will base evidence of asymmetry on p<0.10, and presented intercepts with 90% CIs. Other reporting biases, including selective non-reporting of an outcome, will be evaluated through discrepancies between the registered protocol and the final publication. If we could not find the record of a study in clinical trial registries, we will contact the authors for more information.

### STRATEGY FOR DATA SYNTHESIS

We will only conduct meta-analysis if the included studies are sufficiently homogeneous in terms of design, population, interventions and comparators reporting the same outcome measures.

The results for clinically homogeneous studies will be meta-analyzed using RevMan 5 (50), with the inverse variance method with random effects model. Separate meta-analyses will be presented for specific populations or interventions if statistically significant heterogeneity is explained by some of these, or if a convincing subgroup effect was found.

For any outcomes where insufficient data is found for a meta-analysis, a narrative synthesis will be presented.

### SUBGROUP ANALYSIS AND INVESTIGATION OF HETEROGENEITY

We will conduct our main analyses combining all the interventions, but when possible we will also conduct the following subgroup analyses:

- Patients’ age (< 65 years versus ≥ 65 years).
- Gender (male versus female).
- Duration of rheumatoid arthritis (≤ 1 year versus > 1 year).
- Rheumatoid arthritis disease activity (e.g. remission/low disease activity versus moderate/high disease activity).
- Intervention category (herbal cannabis, cannabis-derived products and synthetic cannabinoids).
- Individual intervention (i.e. specific drugs).
- Route of administration (e.g. inhaled).

If we identify significant differences between subgroups (test for interaction <0.05) we will report the results of individual subgroups separately.

If an unexpected clinical or methodological heterogeneity is found due to obvious reasons, we will state hypotheses regarding these for future versions of this review. We will not anticipate undertaking additional analyses in this version.

### SENSITIVITY ANALYSIS

We will use sensitivity analyses to assess the impact on the treatment effects of inclusion of trials with ‘high risk of bias’ versus ‘low overall risk of bias’ for each comparison and outcome. If the primary analysis effect estimates and the sensitivity analysis effect estimates significantly differ, we will either present the low risk of bias — adjusted sensitivity analysis estimates — or report the primary analysis estimates downgrading the certainty of the evidence due to risk of bias.

If we find substantial clinical heterogeneity on how the condition (rheumatoid arthritis) was defined we will explore this using sensitivity analysis.

### ASSESSMENT OF CERTAINTY OF EVIDENCE

The certainty of the evidence for all outcomes will be judged using the Grading of Recommendations Assessment, Development and Evaluation working group methodology (GRADE Working Group) (53).The certainty of the evidence will be assessed across the domains of risk of bias, consistency, directness, precision and reporting bias. Certainty will be adjudicated as high, moderate, low or very low.

For the main comparisons and outcomes, we will prepare Summary of Findings (SoF) tables and also interactive Summary of Findings (iSoF) tables (http://isof.epistemonikos.org/). A SoF table with all the comparisons and outcomes will be presented as an appendix.

## Data Availability

All data related to the project will be available. Epistemonikos Foundation will grant access to data.

## NOTES

### AUTHOR APPROVAL

This manuscript has been seen and approved by all listed authors.

### CONTRIBUTIONS OF AUTHORS

GR is the guarantor. CS, GR and RB drafted the manuscript with the contribution of all authors. The search strategy was created by CS and MM, a Health Sciences Librarian with expertise in systematic review searching. JD, as a rheumatologist, provided expert advice. All authors read, provided feedback and approved the final manuscript.

### ETHICS

As researchers will not access information that could lead to the identification of an individual participant, the Scientific Ethics Committee of the Pontificia Universidad Católica de Chile granted ethical exemption for the realization of this study.

### PROVENANCE AND PEER REVIEW

Not commissioned.

### DISSEMINATION

Results of this review will be widely disseminated via peer-reviewed publications, social networks and traditional media and will be sent to relevant international organizations discussing this topic.

